# Considerations for Developing and Implementing an Online Community-Based Exercise Intervention for Adults Living with HIV: a qualitative study

**DOI:** 10.1101/2021.11.14.21266319

**Authors:** Bernice Lau, Isha Sharma, Sukhbir Manku, Julia Kobylianski, Li Yin Wong, Francisco Ibáñez-Carrasco, Soo Chan Carusone, Kelly K. O’Brien

## Abstract

**Objectives:** To describe the need for and utility of online community-based exercise (CBE) interventions with adults living with HIV and identify factors to consider in developing and implementing an online CBE intervention with adults living with HIV.

**Design:** Qualitative descriptive study using web-based semi-structured interviews.

**Participants:** We recruited adults representing at least one of five stakeholder groups with experience in CBE and/or HIV: 1) adults living with HIV, 2) rehabilitation professionals, 3) fitness personnel, 4) educators with eLearning experience, and 5) representatives from HIV community-based organizations (CBOs).

**Data Collection:** We asked participants to describe their experiences with online CBE, need and utility for online CBE, and factors in developing and implementing online CBE interventions. We analyzed data using group-based content analytical techniques.

**Results:** Among the 11 participants, most had experience working with adults living with HIV (73%) or with tele-health/rehabilitation/coaching in HIV or other chronic conditions (91%). Participants identified the need and utility for online CBE interventions to increase accessibility and continuity of care with adults living with HIV. Six factors to consider in developing and implementing online CBE included: 1)person-specific considerations (episodic nature of HIV, stigma, HIV disclosure), 2)accessibility of program (physical space to exercise, reliable internet, access to devices, digital literacy), 3)program delivery and technology (live versus pre-recorded online classes, multiple online platforms for delivery, physical activity tracking, troubleshooting technology), 4)attributes of program personnel (working with CBOs, relatable instructors, diverse staff), 5)program content and design (tailored exercise classes, educational sessions) and 6)building community (shared experiences, peer support, social opportunities).

**Conclusions:** There is a need and utility for online CBE in the context of HIV. Considerations for development and implementation span individual, structural and technical, and community dimensions. Results can inform the future development and implementation of online CBE with adults living with HIV and other chronic episodic conditions.

**STRENGTHS & LIMITATIONS OF THIS STUDY:**

- To our knowledge, this is the first qualitative study exploring the perspectives of key stakeholders in developing and implementing an online community-based exercise (CBE) program for adults living with HIV.
- Insights gained from this study can help to inform future online community-based exercise programming for adults living with HIV and other chronic and episodic conditions.
- We used the Model for ASsessment of Telemedicine applications (MAST) framework as a conceptual foundation to design the interview guide, which enabled us to comprehensively explore diverse aspects of telemedicine services in relation to information, communication and technology.
- All interviews were conducted online with participants who had access to technological devices and reliable internet, therefore, results may not be transferable to individuals or contexts without access to technology.
- Most participants were living in Canada, hence, it is unclear how results may be transferable to other geographical contexts.

## INTRODUCTION

The advent of antiretroviral therapy (ART) has increased the longevity of people living with HIV (PLWH).^1^ Nevertheless, the combination of HIV, side-effects of ART, aging, and multimorbidity in PLWH can contribute to multiple complex and episodic physical, cognitive, mental, emotional, and social health challenges, which can be conceptualized as disability.^2-4^ These challenges, such as pain, fatigue, reduced day-to-day activities, anxiety, depression, social exclusion, financial insecurity, and uncertainty regarding future health are associated with disability, poorer adherence to ART, and complexity of health management.^5-7^ Despite these multilevel health challenges, PLWH have demonstrated a capacity for resilience and advocacy from within their community, and readiness to engage in behaviours to improve health outcomes, such as exercise.^8-11^

Aerobic and resistive exercise, particularly a combination, is an effective rehabilitation strategy that can reduce disability and improve health outcomes among PLWH.^12-15^ However, despite the benefits, results from a systematic review indicated 51% of PLWH met the recommended 150 minutes of moderate-vigorous aerobic physical activity per week.^16^ Potential barriers to exercise may include stigma, discomfort with exercise environments, lower socioeconomic status, transportation accessibility, physical symptoms, and the episodic nature of HIV, highlighting the need to consider different models of exercise implementation for PLWH.^17-19^ Community-based exercise (CBE) includes an individual, or a group of individuals with related conditions, engaging in an organized set of exercises under the supervision of a healthcare practitioner or exercise specialist, such as a physical therapist or a fitness instructor in a community-based facility.^20 21^ CBE has been well established and researched in other populations of individuals living with chronic conditions.^22-25^ CBE can cultivate social interaction, encourage regular exercise and facilitate self-management strategies in PLWH to independently manage health challenges.^26-29^ Additionally, CBE can provide physical health benefits in PLWH such as a decrease in lipid profile, waist circumference, diastolic and systolic blood pressure, and an increase in upper and lower body muscular strength.^28 30 31^ Despite the benefits of CBE, PLWH may experience barriers to exercising in traditional gym environments, including, interpersonal, financial, and geographical barriers, self-image issues and stigma, and difficulty initiating exercise following periods of inactivity.^26 29^ Furthermore, events of the COVID-19 pandemic such has gym closures has highlighted the need to consider novel ways in which to facilitate engagement in CBE among PLWH.^32^

Tele-rehabilitation is the delivery of rehabilitation programs or services via technology, such as phone and video calls, web-based platforms, or mobile apps; used initially to increase efficiency, quality of care, and access to healthcare for populations with geographical barriers.^33 34^ This method of delivery has become essential for delivering care in the context of the COVID-19 pandemic with the closure of many in-person services.^35 36^ Tele-rehabilitation can mitigate transportation, financial, and time commitment barriers to CBE, with the potential to improve access to exercise programs for PLWH.^37^ Tele-coaching, a component of tele-rehabilitation that involves the use of technology for remote supervision, guidance, and communication of an exercise program, can help to improve physical activity levels and manage symptoms for people living with chronic diseases.^38-40^ Hence, tele-coaching may be an ideal model to enhance engagement in CBE for PLWH.^41-43^ However, the translation of online tele-coaching CBE programs within an HIV context, and the specific considerations and recommendations for developing and implementing an online CBE program with PLWH is unknown. Our aim was to describe considerations for developing and implementing an online CBE intervention with adults living with HIV, from the perspectives of stakeholders with a role in online CBE implementation with adults living with HIV including: persons living with HIV, rehabilitation or other healthcare professionals, fitness professionals, eLearning educators, and representatives from HIV community-based organizations (CBOs). Specific objectives were: 1) to describe the need for, and utility of, online CBE interventions with adults living with HIV and 2) to identify factors to consider in developing and implementing an online CBE intervention with adults living with HIV.

## METHODS

### Study Design

We conducted a cross-sectional qualitative descriptive study using web-based semi-structured interviews with multiple stakeholders to explore perspectives of online CBE and the HIV community.^44^ This study was approved by the University of Toronto Research Ethics Board (Protocol #39603).

### Patient and public involvement

We consulted with two community members living with HIV with expertise in HIV and exercise for guidance throughout the study. Community members participated in pilot interviews and advised on the development of and provided feedback on the interview guide and process. This research evolved from a long-standing community-academic-clinical partnership among people ageing with HIV, researchers, and clinicians who identified key research priorities in HIV, ageing and rehabilitation as part of the Canada-International HIV and Rehabilitation Research Collaborative (CIHRRC).^45^ Results from this study will be translated via a study summary emailed to study participants and presentations at conferences and community organizations.

### Participants

We recruited ‘online CBE stakeholders’ defined as having a role in online CBE implementation with adults living with HIV, specifically possessing experience, expertise, or interest in areas including any combination of HIV, tele-coaching (exercise), tele-rehabilitation, or eLearning. We recruited adults (18 years of age or older) who self-identified as representing at least one of five stakeholder groups: 1) persons living with HIV, with experience or interest in online exercise applications or interventions (PLWH); 2) rehabilitation professionals or other health care professionals with a role in rehabilitation with people living with chronic disease and experience in tele-health or tele-rehabilitation interventions (RP); 3) fitness personnel or managers engaged in online exercise personal instruction or online exercise class delivery (for-profit and non-profit sectors) (FP); 4) educators with experience in eLearning in the field of rehabilitation or chronic disease management (eL); and/or 5) representatives from community-based organizations (CBOs) with experience delivering health or social support services remotely with PLWH (CBO).

### Recruitment

We purposively identified individuals in the community who possessed experience in at least one of the five stakeholder groups. Members of the team (BL, IS, SM, JK, LYW) sent an email to stakeholders to introduce themselves as MScPT students and to explore interest and eligibility to participate in the study. Stakeholders who responded with interest were scheduled for an interview. We obtained written or verbal consent from all participants prior to participation in the study.

### Data Collection

Two MScPT student team members (one interviewer and one field note taker) conducted interviews with each participant using web-based (Zoom) software.^46 47^ Team members conducting the interviews did not have a prior relationship with participants.

#### Interview guide

We used a semi-structured interview guide tailored for each stakeholder group (Supplemental file 1). We developed the interview guide using the Model for ASsessment of Telemedicine applications (MAST) framework, used to inform the assessment of existing telemedicine applications.^48^ Our interview guide included questions that pertain to each of the eight domains of the MAST framework: health; clinical impact; characteristics of the application; safety; person-specific and environmental-factors; sociocultural, ethical, and legal factors; economic factors and organizational factors.^48^ Based on feedback from two pilot interviews with PLWH and eLearning stakeholders, we developed a PowerPoint slideshow to accompany the interview questions that we shared on screen via Zoom during the interview to facilitate understanding in subsequent interviews (Supplemental file 2). We refined the interview guide twice throughout our study to clarify the probing questions and terminology.

#### Interviews

At the start of each interview, we asked five demographic questions pertaining to age, gender, stakeholder group(s), experiences working with PLWH, and experiences with tele-health/tele-rehabilitation/tele-coaching/exercise online with PLWH or other chronic conditions. We tailored our questions according to the stakeholder group each participant self-identified with during the interview. In the interviews, we asked participants about their perspectives on the need for and utility of online CBE programs with PLWH, the factors important for developing and implementing online CBE interventions with PLWH, followed by overall recommendations for developing and implementing an online CBE program in the context of HIV. We defined the ‘need’ for online CBE programming as filling or addressing a gap in health care and ‘utility’ as how this intervention could be useful with PLWH. Interviews were recorded on an audio recorder. Participants received a $30 electronic gift card as a token of appreciation for their participation in the study. All interviews were audio recorded using a digital recorder and later transcribed verbatim.

### Data Analysis

#### Interview Data

We performed data collection and analysis simultaneously. We reviewed the transcripts for accuracy. We used a combination of Braun and Clarke’s thematic analysis procedure with the DEPICT group-based approach to inform our data analysis.^49 50^ Our group-based analytical approach consisted of six steps: 1) dynamic reading (all students [BL, IS, SM, JK, LYW] and at least one advisor [FIC, SCC, KKO] reviewed the first five transcripts to note first impressions of the data and create initial codes relating to the study objectives), 2) codebook development (students met as a group to discuss agreed upon codes and establish a common technique for coding future transcripts and generation of a codebook), 3) coding remaining transcripts (the remaining six transcripts were divided equally among three students [IS, SM, LYW] and coded independently) 4) inclusive reviewing and summarizing of categories (codes were organized into categories and summaries developed for each category), 5) collaborative analyzing (students met as a group to make sense of data and create a figure to illustrate findings), 6) translating (students and advisors met throughout to refine the results for knowledge translation). We used NVivo software to facilitate data management (QSR International, NVivo, V.12)^51^ and developed participant summaries that included demographic information.

#### Demographic Data

We calculated medians and 25-75^th^ percentiles for continuous variables (age) and frequency (%) for categorical variables (gender; stakeholder group; experiences working with PLWH; delivering online CBE).

### Sample Size

We used a combination of purposive and snowball sampling, with the aim of recruiting a sample of 12 to 15 participants, with ≥2 participants from each of the five stakeholder groups.^52^ Previous studies using qualitative descriptive methods to explore (in-person) CBE for PLWH used a similar sample size, with three stakeholder groups from which they were able to recruit a minimum of two participants each.^27^ Based on previous studies, we did not anticipate this proposed sample size to reach saturation. However, this was not the goal of our study as our aim was to recruit a sample that enabled us to capture a diversity of perspectives and experiences in the context of online CBE interventions, allowing us to adequately address study objectives.

## RESULTS

Of the 24 CBE stakeholders approached, 11 participated in an interview between January and May 2021, lasting between 60-90 minutes in duration. The median age of the participants was 49 years (25-75^th^ percentile: 40, 58); and the majority (73%) identified as women and lived in Canada (91%). The majority were rehabilitation professionals (63%). Eight (73%) participants had prior experience working with PLWH and almost all participants (91%) had experience with tele-health, tele-rehabilitation, tele-coaching, or online exercise for PLWH or other chronic conditions (Table 1). Of the 11 participants, 10 (91%) represented two or more stakeholder groups.

**Table 1.**
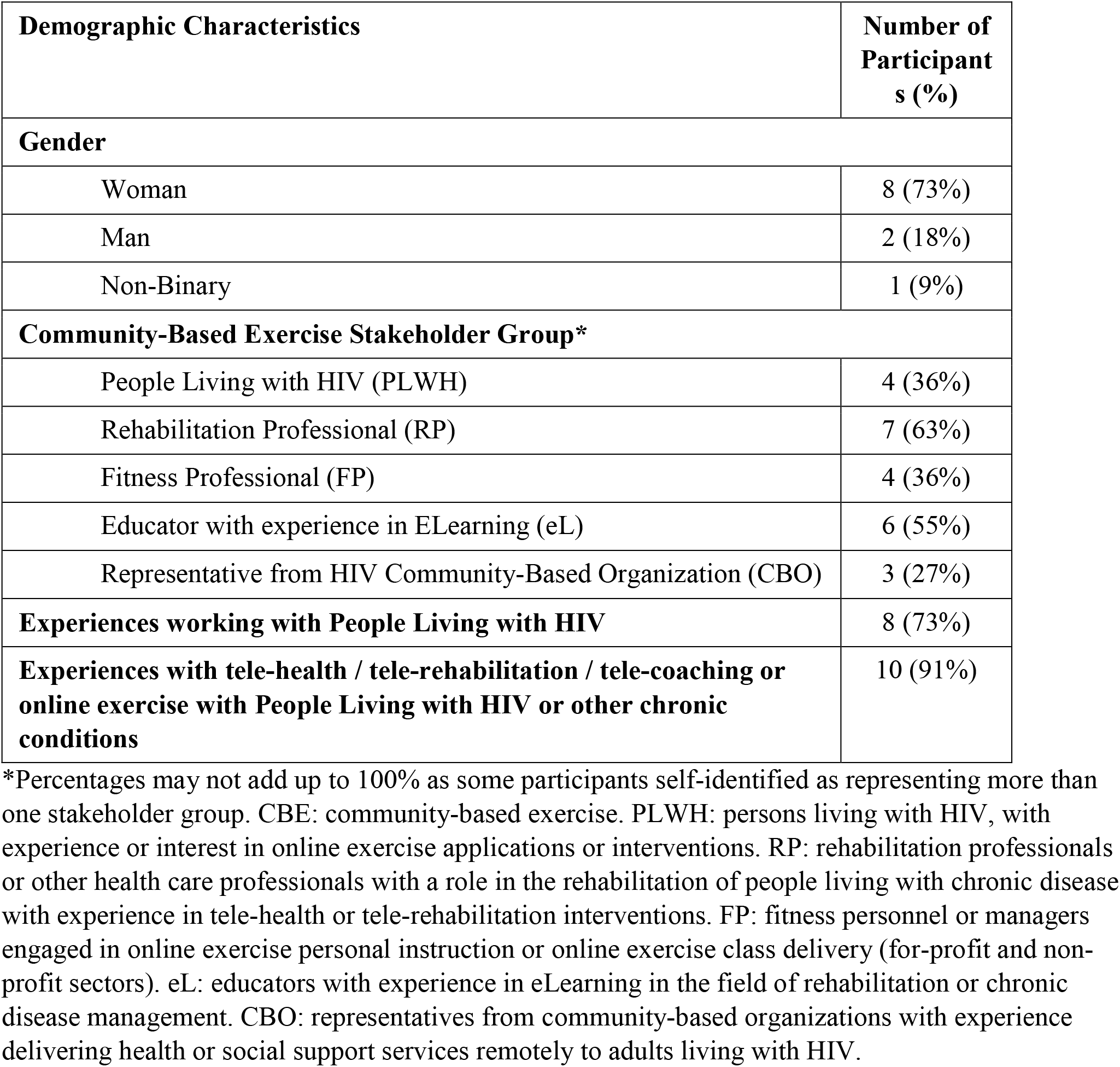
Characteristics of Participants (n=11)

| **Demographic Characteristics** | **Number of Participants (%)** |
| --- | --- |
| **Gender** |  |
| Woman | 8 (73%) |
| Man | 2 (18%) |
| Non-Binary | 1 (9%) |
| **Community-Based Exercise Stakeholder Group\*** |  |
| People Living with HIV (PLWH) | 4 (36%) |
| Rehabilitation Professional (RP) | 7 (63%) |
| Fitness Professional (FP) | 4 (36%) |
| Educator with experience in ELearning (eL) | 6 (55%) |
| Representative from HIV Community-Based Organization (CBO) | 3 (27%) |
| **Experiences working with People Living with HIV** | 8 (73%) |
| **Experiences with tele-health / tele-rehabilitation / tele-coaching or online exercise with People Living with HIV or other chronic conditions** | 10 (91%) |

\*Percentages may not add up to 100% as some participants self-identified as representing more than one stakeholder group. CBE: community-based exercise. PLWH: persons living with HIV, with experience or interest in online exercise applications or interventions. RP: rehabilitation professionals or other health care professionals with a role in the rehabilitation of people living with chronic disease with experience in tele-health or tele-rehabilitation interventions. FP: fitness personnel or managers engaged in online exercise personal instruction or online exercise class delivery (for-profit and non-profit sectors). eL: educators with experience in eLearning in the field of rehabilitation or chronic disease management. CBO: representatives from community-based organizations with experience delivering health or social support services remotely to adults living with HIV.

### Need and utility for online CBE interventions in the context of HIV

Participants described the “need” for online CBE and its “utility” as interconnected. These categories represent the need for access to health services by PLWH, especially those aging, who may experience difficulties with self-image, feeling stigmatized (perceived stigma), mobility issues and incidental costs such as membership fees and transportation.

The *need* for online CBE was reported as amplified during the COVID-19 pandemic. Most participants confirmed that online CBE was necessary, beneficial and diversifies options for access by PLWH. This increase in accessibility was described as geographical independence. Not having to travel to attend gym environments to exercise increased the overall accessibility of CBE for populations who may live in more remote areas:

> *“You might not have a role model fitness instructor that’s specialized in HIV. But by moving services online you’ve got a much bigger reach to deliver care and to allow people to connect with others*.” (P04; RP, FP, eL stakeholder).

The *utility* of online CBE to address gaps in accessibility to health services was described as the ability to enhance continuity of care and improve multidimensional health outcomes for people living with HIV. One participant with experience delivering CBE for stroke survivors spoke about the potential for online CBE to provide on-going healthcare management; to:

> *“[expand] the window of opportunity for rehab”* both between healthcare appointments or after leaving a particular setting or program as *“they [end-users] may continue on an outpatient basis for a couple of months, [and] can now be at home and still have access to a therapist, two days a week through our tele-rehab program*.*”* (P01; RP, eL stakeholder).

Notably, participants suggested online CBE may be used to support physical and mental health dimensions of PLWH, used as “*their primary form of exercise*” or have “*specific health needs that they have anxiety around or that may need support with*.” This utility of online CBE was a “*tool in the toolbox*” to support healthy aging in this population. A participant involved in online CBE program delivery for different populations mentioned: “*Just keeping people in good physical and emotional health helps healthy aging*.” (P04; RP, FP, eL stakeholder).

### Factors in developing and implementing an online CBE intervention with adults living with HIV

We identified six factors to consider in developing and implementing online CBE for PLWH, all of which were interconnected across individual, structural and community dimensions (Figure 1).

**Figure 1.**
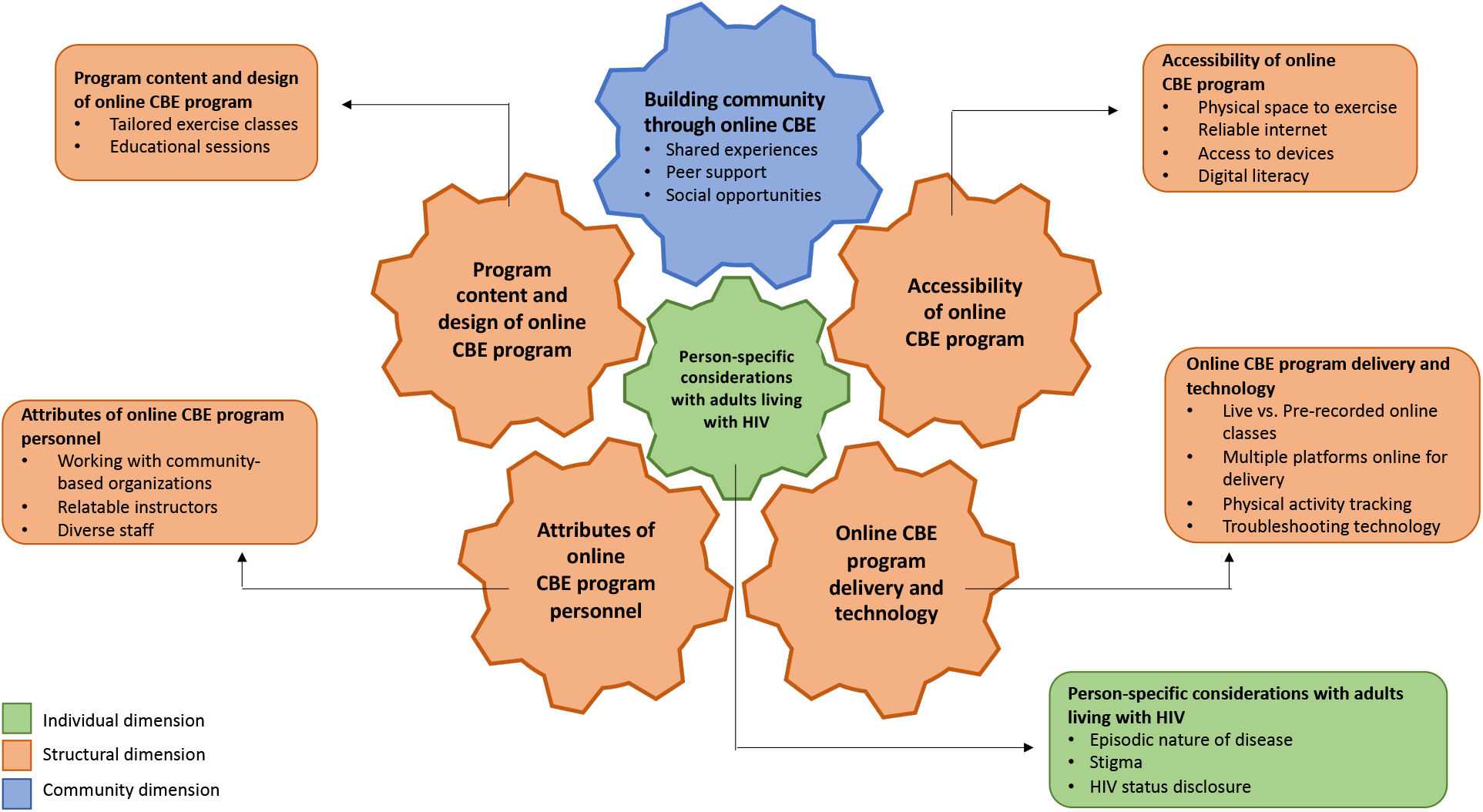
Six factors to consider in developing and implementing an online CBE intervention with adults living with HIV.

At the center was the individual dimension, referring to person-specific considerations with adults living with HIV, which were integral and at the forefront when creating person-centered online CBE programming. Surrounding the individual dimension was the structural dimension, defined as the logistical and technical considerations providing the groundwork for online CBE, and include accessibility of program, program delivery and technology, attributes of program personnel, and program content and design. The individual and structural dimensions were interconnected with the community dimension, consisting of opportunities for building community with online CBE.

#### Individual Dimension

##### Person-Specific considerations with adults living with HIV

Participants mentioned the importance of understanding the complexities of living with HIV as they relate to in-person and online CBE, including challenges with the episodic nature of the disease, stigma, and HIV status disclosure:

> *“HIV is an episodic disability*…*the clients I see are generally quite complex, they have multiple comorbidities or multimorbidity*…*some people say, 80-90% of our clients have mental health issues and almost as high of percentage have substance use issues*.*”* (P03; RP, FP, eL stakeholder).

Participants also discussed the importance of understanding HIV-related stigma and the intersectionality of gender, sexual orientation, and body image, and how it affects access and willingness to exercise, including through an online platform:

> *“I think that there is a lot of fear of being judged, by [*…*] fitness instructors, employees within the gym, other people at the gym. I think generally speaking from my experience, people living with HIV would rather have an HIV only exercise group. But that comes with a confidentiality breach, a potential one*.*”* (P10; RP, FP, eL stakeholder)

As such, when participating in online CBE specifically for PLWH, participants described concerns regarding confidentiality and fear of HIV disclosure when participating in online CBE programming:

> *“you’d hate for someone to be outed as being HIV when they’re not out in the community, or to their friends or to their family*.*”* (P02; PLWH stakeholder).

These potential challenges and barriers should be considered when designing online CBE programs with adults living with HIV.

#### Structural Dimension

##### Accessibility of Program

Participants highlighted the importance of considering the extent of access by PLWH to online CBE programs including having the physical space to exercise, reliable internet, access to devices, such as computers, tablets, smartphones, and the digital literacy skills to use these devices:

> *“The older population may not have Internet or may not know how to use the Internet. Then access to technology, you’d have to take the time to explain Zoom if that’s the platform that you’re going to be using for the program [*…*] So there’s a lot of pre-exercise program technology that needs to be done with individuals*.” (P07; PLWH, eL, CBO stakeholder)

To make online CBE inclusive and accessible, participants suggested offering a modicum of training to PLWH to enhance their digital literacy as described by a participant involved with online CBE for stroke survivors:

> *“We’re using Zoom as the platform [*…*] what we’re trying to do to help support people is that they have some, ‘virtual intake sessions’ so that we have some time in those sessions to actually talk them through: How do you navigate Zoom? We have a manual that we sent to them and they can install it on their device ahead of time, and we’re there to troubleshoot*.” (P01; RP, eL stakeholder)

##### Program Delivery and Technology

All participants discussed CBE program delivery and its associated technology as an important factor, which included considerations for live versus pre-recorded online exercise classes, using multiple online platforms for delivery, physical activity tracking, and troubleshooting technology. Participants noted the importance of using synchronous and asynchronous modalities to deliver online CBE such as pre-recorded classes, which were advantageous for those “*who want to exercise in their own time or those who cannot join them live*”, meanwhile live classes would allow for instructor real-time correction and guidance as well as interaction among end-users.

Participants noted the importance of knowing how to use multiple platforms, such as Zoom and Microsoft Teams, to meet the needs of end-users. Notably, Zoom was the most mentioned platform used by participants. While some participants expressed concerns regarding privacy and usage of tracking data, options to track physical activity were described by others to be potentially beneficial to the end-user:

“*Data tracking for motivation and monitoring is really important ‘cause then you could see how much you’re progressing or where you have to work on something a little bit more*.” (P06; PLWH, CBO stakeholder).

Participants highlighted some challenges created by the implementation of online CBE, specifically referring to troubleshooting technology related to audio/visual components, internet reliability, and devices used for online delivery of CBE programs:

> *We need a strong Wi-Fi, and we need a nice mic - nicely set, then we can deliver the voice, the music at once. So, if one of those [things has] small hiccups, the experience is not going to the members*. (P05; RP, FP stakeholder)

##### Attributes of Program Personnel

Participants involved in previous online CBE programming reported the importance of having relatable and diverse staff who are able to liaise and work with community-based organizations, especially HIV community based organizations (CBOs). One participant with experience using online CBE suggested: *“some of the best resources are probably your local AIDS service organizations*”. Having a relatable instructor also was discussed as an important factor by many participants, as:

“*they [end-users] also identified with her [the instructor] in a way because she also was coming from trauma*.” (P06; PLWH, CBO stakeholder).

Finally, a participant involved in the development of an online exercise platform for individuals living with chronic conditions discussed working with clinicians, charities and academics to increase CBE program awareness. They mentioned how forming a “multidisciplinary team” includes:

> *“[a] technical team or user experience [personnel], data analytics… tech support, customer service, studio managers, clinical leads, instructors, marketing, developers, user experience design and partners*.” (P04; RP, FP, eL stakeholder)

##### Program Content & Design

Participants highlighted the importance of supporting the needs of PLWH in online CBE, including offering tailored exercise classes and educational sessions focused on enhancing multidimensional health and self-management among PLWH. Commonly reported components to include in an online CBE program were exercise classes tailored to the physical ability of end-users:

“*not just offering one type of intervention exercise program, recognizing that people have different needs*,” P04 (RP, FP, eL stakeholder).

Educational sessions for end-users on HIV and health management also were described as desirable by participants with experience delivering online CBE. Topics covered through these sessions included education on the “*benefits of exercise*,” nutrition, “*like what kind of nutrition can help you maintain an exercise routine*,” mental health considerations such as “*[lectures on] sleep hygiene… meditation… and stress management”* as well as what to be aware of when taking certain medications.

#### Community Dimension

##### Building Community

Building a sense of community when creating an online CBE program was discussed as an important factor in online CBE by all participants; highlighting the value of end-users connecting to one another through shared experiences, peer support, and social opportunities. One participant mentioned online exercise could allow for:

> *“room for them to share experiences or just how they’re feeling in general, their aches and pains that they can each relate with*…*it might be like a factor for them to be motivated because they find that people here understand me and what I’m going through*.” (P11; RP, eL stakeholder).

Another participant, with experience as an end-user of online CBE, highlighted feelings of empowerment nurtured by a program with a group of people with similar experiences:

> *We can try to chat privately if it’s on, Zoom and say hey, ‘here’s my phone number let’s talk*.*’ [*…*] you could actually gain friends and I have found that, when people living with HIV come together, there is power and support*. (P07; PLWH, eL, CBO stakeholder)

Forming these connections can facilitate peer support, which can increase physical activity and exercise program adherence as on participant stated:

“*they’ll [end-users] form relationships, they’ll continue with long term engagement in physical activity, and supporting and encouraging one another*.” (P04; RP, FP, eL stakeholder)

However, some participants reported that community building online differs from in-person CBE:

“*some of that kind of informal chatter and conversation, those opportunities are lost, so we may have to rethink, how do we still get to the best parts of in-person interactions and translate it into a virtual format*.*”* (P01; RP, eL stakeholder)

To address this, participants suggested including designated time before and/or after an online CBE class to socialize, “*whether that’s through a group chat with everyone participating together or you know, just being online for another 15 minutes after the exercises to check on each other*.” (P11; RP, eL stakeholder)

## DISCUSSION

To our knowledge, this is the first qualitative study to explore considerations for developing and implementing an online CBE program with PLWH. Results suggest there is a need for, and utility of, online CBE with PLWH from both organizational and end-user stakeholder perspectives. Participants revealed six interconnected factors to consider, spanning individual, structural and community dimensions, when developing and implementing such a program.

### Need for and utility of online community-based exercise with people living with HIV

Participants highlighted the need for and utility of online CBE to increase access to health services by enhancing continuity of care and improving multidimensional health outcomes for PLWH. In previous studies, barriers to CBE and other forms of exercise for PLWH included travelling through cold weather and costs of a gym membership.^26 27 29^ Similarly, individuals living with COPD expressed long travel times as limiting their participation in exercise,^53^ and those living in rural communities described difficulties accessing health services.^54^ Participants in this study articulated how an online CBE intervention may be a valuable tool for adults living with HIV or other chronic conditions, especially if end-users do not need to travel to access an exercise program.

Notably, participants emphasized how online CBE may be uniquely poised to fill health care delivery gaps for PLWH. Pilot studies investigating existing mobile health technologies for PLWH focused on physical health, but have limited mention of supporting other health dimensions.^41^ Participants in this study described online CBE as a means of improving health in the physical, mental, and social dimensions; with social health strongly linked to the factor of building a sense of community. This is important, as creating community, leadership, and resiliency are strengths within the HIV community.^10 11^ Thus, creation of online CBE should include engagement of PLWH at all stages of development and implementation.

### Navigating person-specific considerations with adults living with HIV

Persons living with HIV can experience high levels of perceived, anticipated and enacted stigma,^55 56^ and a barrier to in-person exercise, which may increase social isolation for PLWH.^18 27^ As group-based forms of CBE can provide opportunities for social interaction, PLWH participating in an HIV-specific online CBE intervention may feel more comfortable participating with other PLWH who possess a mutual and shared understanding and shared experiences living with HIV.^57^ Hence, an online CBE program designed exclusively for PLWH may be preferred by some PLWH. However, a PLWH-specific online CBE program may pose additional confidentiality and privacy challenges, as users of online exercise programs often display their name and photo or video during exercise sessions. Thus, participation in an HIV-specific program may lead to unintentional disclosure of HIV status. HIV disclosure could be mitigated through a generalized online CBE for individuals living with chronic conditions, including PLWH. Ultimately, benefits exist with both HIV-specific and generalized chronic (episodic) CBE programming depending on the individual.

### Addressing digital literacy

Participants in this study shared concerns regarding the usability of online CBE programs for those with limited computer skills. This was supported by rehabilitation professionals who suggested there may be unequal access to remote health services based on end-users’ level of digital literacy.^53 54^ However, the prospect of learning new technologies and information technology (IT) skills has been met with resistance from rehabilitation professionals who stated that helping end-users with technological troubleshooting was not within their scope of practice.^53^ Within the context of the COVID-19 pandemic, this mindset may have shifted as the increased need for virtual care may encourage professionals to adopt tele-rehabilitation and increasing willingness to embrace the use of IT.^58^ Nonetheless, a multidisciplinary team (including those with knowledge of IT) are needed for successful implementation of online CBE.

### Forming social connections in online CBE settings

Participants in this study indicated that if PLWH participate in a class with others who share similar experiences, it can encourage the formation of social connections. This is important as social isolation, particularly during the COVID-19 can be a contributor to poorer health outcomes and difficulty engaging in exercise for PLWH.^59 60^ Participation in an online CBE intervention can optimize community building and help to decrease social isolation and increase engagement in exercise among people with other chronic conditions, thus potentially possessing transferability with PLWH.^61 62^ However, challenges in forming social connections online may persist as some aspects of socialization may not occur naturally in online settings, such as the informal chatter and conversations mentioned by participants. To help facilitate such interactions, previous studies examining online exercise programs demonstrated the necessity of providing opportunities for “icebreaking” pre- and post-class.^53^ Thus, we recommend including designated social time for end-users to interact with one another as well as with instructors during online CBE programming.

### COVID-19 pandemic context

This study was designed prior to the COVID-19 pandemic as a way to address barriers to exercising in traditional gym environments among PLWH. Due to the timeline of the pandemic, we collected data at a time when participants were having to shift from in-person to online services out of necessity and fitness facility closures. As a result, the need for online CBE programs and how they may be used, were often interwoven with personal and professional experiences from the COVID-19 pandemic; including conversations around how online services and programs may be needed, have benefit for, and/or be used, following the pandemic. With the closure of fitness facilities during the COVID-19 pandemic and the potential reluctance for PLWH to return to gym/fitness facilities in-person upon reopening, these results may also have additional relevance and meaning to PLWH as well as those living with other chronic or episodic conditions.

### Implications for future research

Our results contribute to the understanding of considerations for developing and implementing an online CBE intervention with adults living with HIV, from the perspectives of five stakeholder groups. Using the six factors for consideration in this study, future work may involve the development of an online CBE intervention in collaboration with the HIV and rehabilitation communities. Future research should examine the strengths, challenges and impact of implementation of online CBE in ‘real world’ community settings from the perspectives of PLWH and individuals involved in administering the CBE. Evaluation of the impact of online CBE on physical, mental, and social health-related outcomes following an online CBE intervention, as well as research into the cost-effectiveness and sustainability of an online CBE intervention is needed.

### Strengths and Limitations

Our study included perspectives from participants with lived experience, expertise in eLearning and tele-heath/tele-rehabilitation in the context of chronic disease management, allowing for a diversity of insights into online CBE and chronic and episodic conditions such as HIV. Our use of the MAST framework enabled us to comprehensively explore the diverse aspects of telemedicine services using this conceptual foundation of information, communication and technology.^48^ We did not achieve our targeted sample size of 12 participants, nor did we reach saturation. Nonetheless, we were able to capture a rich description of experiences and diversity of perspectives enabling us to achieve our study objectives. Conducting interviews online using Zoom implied participants had access to technology and the internet. As a result, perspectives may not be transferable to those without access to technology and who may have articulated financial, technical and digital barriers for online CBE. Further, most participants were living in Canada, hence, it is unclear how results may be transferable to other geographical contexts.

## CONCLUSIONS

We described the need and utility for online CBE among PLWH and six factors spanning individual, structural and community dimensions for considerations when developing and implementing online CBE with PLWH. Results provide a foundation for the future development and implementation of online CBE programs for PLWH and may help to inform online CBE development for individuals living with other chronic and episodic conditions.

## Data Availability

All data produced in the present study are available upon reasonable request to the authors

## ACKNOWELDGEENTS

This research was completed in partial fulfillment of the requirements for a Masters in Physical Therapy degree at the University of Toronto. The authors thank the participants for their time and sharing their insights. We also thank Francisco Ibáñez-Carrasco and George Da Silva, for taking part in pilot interviews and providing feedback on the interview guide.

## CONTRIBUTORS

KKO, SCC and FIC designed the study and provided guidance throughout the research process. KKO, SCC and FIC possess expertise in qualitative methodology and HIV and exercise research. KKO, SCC and FIC supervised BL, IS, SM, JK and LYW (MScPT students) who developed the protocol, collected and analyzed data and drafted the manuscript in partial fulfilment of the requirements for a MScPT degree at the University of Toronto. BL, IS, SM, JK and LYW developed skills in qualitative methodology and study design; understanding steps of recruitment, data collection and analysis; completing a literature review; developing the research protocol, interview guide, demographic questionnaire, coding scheme and considering ethical issues associated with this research. All steps were closely reviewed and guided by KKO, SCC and FIC. All authors read and approved the final manuscript.

## ETHICS APPROVAL

This study received approval from the University of Toronto Research Ethics Board (Protocol #39603).

## DATA SHARING STATEMENT

Relevant data to this study are included in the article. No additional data is available in accordance with our protocol approved by the University of Toronto Research Ethics Board.

## SUPPLEMENTAL FILES

**Supplemental file 1**: Semi-structured interview guide.

**Supplemental file 2**: PowerPoint slideshow with interview guide questions shared with study participants during Zoom interviews.

**Supplemental file 3**: Original Protocol – Research Ethics Board Approval

