## Supplemental File 1 for "Considerations for Developing and Implementing an Online Community-Based Exercise Intervention for Adults Living with HIV: a qualitative study"

#### Supplemental File 1 - Interview Guide

### Tele-Coaching in HIV and Exercise: Considerations for Developing and Implementing an Online Community-Based Exercise (CBE) Intervention for Adults Living with HIV

##### Preamble:

Thank you for agreeing to participate in this study. My name is ________, and this is _______. We are students in the physiotherapy program at the University of Toronto. As you know, we are interested in your experiences, insights and perspectives on online forms of community-based exercise interventions (or tele-coaching) for persons living HIV. We are conducting this research study to identify factors to consider when developing and implementing an online community-based exercise intervention for people living with HIV from the perspectives of key stakeholders with roles in community-based exercise.

I will be conducting the interview and _______ will take notes to help us better understand the points raised in the interviews. Have you read and are you in agreement what is involved in the study? Do you consent to continue with the interview?

I will be asking a series of questions about your perspectives on, community-based exercise programs, use of technology in healthcare, and how this can be incorporated in developing an online community-based exercise intervention for persons living with HIV.

In this interview I will ask you general questions about your experiences and perspectives. There are no right or wrong answers. Feel free to interrupt me to ask questions or clarifications, skip questions, take a break, or stop the interview all together.

Do you have any questions for us before we begin?

Do you mind if we take notes and record the audio during the interview?

**start recording**

##### Demographic Data

Before we begin, I have a few brief demographic questions. The answers to these questions will help us describe (in general) the characteristics of the participants who took part in the study

| Question | Response options |
|---|---|
| 1) What is your age? In years | ________________ years |
| 2) What gender do you identify with? | ❑ Woman<br>❑ Man<br>❑ Trans: Man to woman<br>❑ Trans: Woman to man<br>❑ Non-binary<br>❑ Two-spirit<br>❑ Other – Please describe: _________ |

| | |
|---|---|
| 3) As you know, we are trying to gather perspectives from different stakeholders to inform recommendation for online CBE with people living with HIV. Please indicate (yes or no) whether you identify with each of the following stakeholders. | a.) Person living with HIV with experience or interest in online exercise applications or interventions<br>❑ Yes<br>❑ No<br>b.) Rehabilitation professionals or other health care professionals with a role in the rehabilitation of people living with chronic disease with experience in tele-health or rehabilitation interventions<br>❑ Yes<br>❑ No<br>c.) Fitness personnel (trainer, manager, coach etc.) engaged in online exercise personal instruction or online exercise class delivery (for-profit and non-profit sectors)<br>❑ Yes<br>❑ No<br>d.) Educators with experience with eLearning in the field of rehabilitation or chronic disease management<br>❑ Yes<br>❑ No<br>e.) Representatives from AIDS Service Organizations (ASO), with experience delivering health or social support services remotely<br>❑ Yes<br>❑ No |
| 4) Experience working with persons living with HIV? | ❑ Yes<br>❑ No |
| 5) Experience with Tele-Health/ Tele-Rehabilitation/Tele-coaching/Exercise online for persons living with HIV or other chronic conditions? | ❑ Yes<br>❑ No |

#### INTRODUCTION

Can you tell me about your experiences with or interest in online tele-health/ tele-rehab / or tele-coaching/online exercise with PLWH or other chronic conditions?

(*If there is an opportunity, interviewers may wish to ask about anecdotal story...*)

**Section 1: NEEDS & UTILITY**

1) Given your experiences, what do you think might be some of the barriers faced by persons with HIV experience when it comes to engaging in regular exercise?
    a. *Possible barriers to probe may include:* physical symptoms, mental health, access, social factors (stigma, SES, lack of inclusion), perception of physical activity, effect of COVID-19 on these factors (social isolation, no exercise programming), general access to facilities, stigma, body image, unique barriers faced by trans community with traditional cis gendered change rooms in gyms, etc.
    b. *Possible considerations:* motivation, readiness to exercise, needs for extra support

**Our study focuses on community-based exercise delivered online. Community-Based exercise is defined as "a group of people with similar conditions exercising together by an organized set of exercises under the supervision of a healthcare practitioner." We know that this form of exercise is one of the tools used to help people living with HIV manage their treatment and overall health:**

2) Can you tell us about your experience or knowledge with respect to any current programs which fall under Community Based Exercise (CBE in general, not only online)?
    a. Can you describe the more...
        i. Successful aspects of these programs?
        ii. Unsuccessful aspects of these programs?
3) How can CBE be used to overcome barriers to exercise for PLWH?
4) What do you think are some facilitators to engaging in CBE for PLWH?
5) What are some of the barriers to engaging in CBE for PLWH?

**The term "Tele-coaching," involves the use of technology for remote supervision, guidance, and communication of a CBE program – in this study we refer to tele-coaching as online CBE (program or intervention), which involves exercising at home under supervision of a fitness coach online or engaging in group-based exercise classes online with an instructor…or following online apps / recorded classes.**

6) Do you think there is a need for ***online*** CBE interventions for PLWH?
    a. If so, can you describe the need(s)?
    b. Do you think that online forms of CBE might facilitate or help overcome some of these barriers (from Q5)? (*And if so:* How?)
7) Do you think there is a use for an online CBE program for PLWH?
    a. What would be the purpose of an online CBE program for PLWH *in the community at large?*
    b. How might an online CBE program be useful for you (or your organization)?
8) Who do you think would most benefit from this type of intervention (online CBE)?
    a. Why do you think they would benefit most?
9) Who do you think may not benefit from (or may not need) this type of intervention?
    a. Why would they not benefit?
    b. What aspects of online CBE might they not need?

#### Section 2: FACTORS

**For this interview, we're going to be using a model called the MAST framework, so that we can touch on all aspects to consider when developing and implementing a tele-coaching-based program.** *Provide document that outlines the frameworks via link through Zoom.* **Your answers to questions throughout the interview might cover topics under more than one domain, but just note that we may still follow up in asking questions particular to each domain to get a deeper understanding of your perspective.**

**Health** + Characteristics of the application

1) Are there any physical, mental health, or social health **barriers** that might affect participation or engagement with online CBE?
    a. **Person living with HIV:**
        i. Are there any physical aspects of HIV, or ART that could affect your willingness to participate or engage in the program?
        ii. Are there any concurrent health conditions that could affect your willingness to participate or engage in the program?
2) What about **facilitators** that might help engagement with online CBE**?**

Clinical impact

3) What are your thoughts on online CBE and its ability to impact disability (I.e. symptom management, wellbeing)? What about healthy aging?

Health + **Characteristics of the application**

4) What are some technical factors that may affect the viability of an online CBE program? (reliability of internet, digital literacy, access to technology)
    a. What are some of the technical problems (use, knowledge, access to smart phone, internet, tablet or computer) we would need to be aware of if we were to implement an online CBE program to different users? PLWH?
5) What content should an online CBE program include, in order to address the needs of clients/patients?
    a. Live vs pre-recorded format
    b. Features of the app/platform
6) What platforms do you use for tele-rehabilitation/tele-coaching/online exercise/online service delivery? (Web-based platforms, teleconferences, smart-phone apps, etc)
    a. What do you like/dislike about these platforms?
7) What are your thoughts regarding data-tracking for motivation and monitoring progress (e.g. using wireless activity monitors like apps or a Fitbit to motivate and measure physical activity)?
    a. Do you think data should be tracked? Why or why not?

Safety

8) Can you speak about methods used by organizations providing the online programs, as well as the users, to ensure privacy and confidentiality?
9) Do users generally feel *physically* safe while using online exercise programs?

Person-specific factors and environmental factors

10) How do you feel end-users **trust** using online platforms for exercise programs?
    a. **Person living with HIV:** 1) How comfortable are you/would you be using technology for exercise programs (specifically for CBE)? *"readiness to participate in online CBE"*
11) Are there any personal or intrinsic factors *(such as disposition, gender, affinity with social cultural or ethno-racialized groups, for example: gay men and gym culture, racism; homophobia, HIV stigma, etc.*) that might affect an end-users **access** to a tele-coaching based program?
    a. Could any of those personal or intrinsic factors affect **adherence** to an online CBE program?
12) We know that the social and physical environment are important to accessing exercise programs, especially considering the recent COVID-19 pandemic. Are there any environmental factors (*potential examples include: internet, digital literacy, ICT devices, physical space at home, equipment, social support, medical clearance to independent exercise) that* may affect **access** to online CBE?
    a. Could any of those environmental factors affect **adherence** to an online CBE program?
13) Can you speak to any social factors that might affect **access** to online CBE? (*such as the potential to interact with others as a part of peer-based online CBE group…for motivation, mutual encouragement, and support*)
    a. Could any of those social factors affect **adherence** to an online CBE program?

Sociocultural, ethical, and legal factors

14) Are end-users able to advocate for themselves if they feel their needs are not being met through an online platform?
15) Can you think of any ethical consequences of widespread implementation of online CBE programs?
    - Would any people/populations be disadvantaged due to this?
16) **Persons Living with HIV**:
    a. Can you speak more about the stigma surrounding HIV and how that might affect your willingness to participate in or engage with an online CBE intervention?

Economic factors

17) How might the cost of participating in/delivering an online CBE program differ compared to in-person programs?
    a. **eLearning experts:**
        i. What are the usual costs of building online-based programs? Apps?
        ii. What's the most expensive process/application for the organization/user? Cheapest? Greatest value in terms of cost/benefit analysis?
18) What is the feasibility of providing CBE online compared to in-person?

Organizational factors

19) In your experience, what is the best way to monitor the success of an online CBE program (or a similar online exercise program)?

20) Can you speak about the culture surrounding the use of online platforms for health services and exercise delivery?
21) Can you speak about your experience (good and bad) during the process of switching from in-person to online services?
22) What resources do you think would be needed for implementation of an online CBE?
    a. Which personnel would be essential for the implementation of an online CBE program?
        i. What would these personnel be responsible for? (I.e. scheduling, intake, coordination, IT support etc.)
        ii. Can you speak to any possible training needed for personnel involved?
    b. What type of education or training is needed for instructors in order to successfully implement online forms of eHealth or CBE delivery? For user?

**Section 3: RECOMMENDATIONS**

We are nearing the end of the interview. We are asking all stakeholders:

23) What do you think are some overall recommendations that will be important for developing and implementing a future online CBE program for PLWH?

**Closing Question and remarks**

24) Is there anything else you would like to tell us regarding online-based services, or any suggestions for implementing an online CBE for people living with HIV?

That brings us to the end of the interview. Thank you for participating, we greatly appreciate you taking the time to share your experiences and perspectives with us. As a token of appreciation, you will receive an attached $30 electronic gift card in your email within the next 7 days.
