## Supplemental File 2 for "Considerations for Developing and Implementing an Online Community-Based Exercise Intervention for Adults Living with HIV: a qualitative study"

### TELE-COACHING IN HIV AND EXERCISE:

### Demographic Questions

1. What is your age in years?

### 2. What gender do you identify with?

- ❑ Cis Woman
- ❑ Cis Man
- ❑ Trans: Man to woman
- ❑ Trans: Woman to man
- ❑ Non-binary
- ❑ Two-spirit
- ❑ Other – Please describe ___________

### 3. Please indicate (yes or no) whether you identify with each of the following stakeholders groups.

a. Person living with HIV with experience or interest in online exercise applications or interventions **(Yes or No)**
b. Rehabilitation professionals or other health care professionals with a role in the rehabilitation of people living with chronic disease with experience in tele-health or rehabilitation interventions **(Yes or No)**
c. Fitness personnel (trainer, manager, coach etc.) engaged in online exercise personal instruction or online exercise class delivery (for-profit and non-profit sectors) **(Yes or No)**
d. Educators with experience with eLearning in the field of rehabilitation or chronic disease management **(Yes or No)**
e. Representatives from AIDS Service Organizations (ASO), with experience delivering health or social support services remotely **(Yes or No)**

4. Experience working with persons living with HIV? **(Yes or No)**

5. Experience with Tele-Health/ Tele- Rehabilitation/Tele-coaching or Exercise online for persons living with HIV or other chronic conditions? (Yes or No)

### Section 1

NEEDS & UTILITY

1. Given your experiences, what do you think might be some of the barriers faced by persons with HIV experience when it comes to engaging in regular exercise?

2. Can you tell us about your experience or knowledge with respect to any current programs which fall under Community Based Exercise (CBE in general, not only online)?

### 3. How can CBE be used to overcome barriers to exercise for PLWH?

4. What do you think are some facilitators to engaging in CBE for PLWH?

5. What are some of the barriers to engaging in CBE for PLWH?

6. Do you think there is a need for online CBE interventions for PLWH?

7. Do you think there is a use for an online CBE program for PLWH?

8. Who do you think would most benefit from this type of intervention (online CBE)?

9. Who do you think may not benefit from (or may not need) this type of intervention?

### Break?

### Section 2

FACTORS

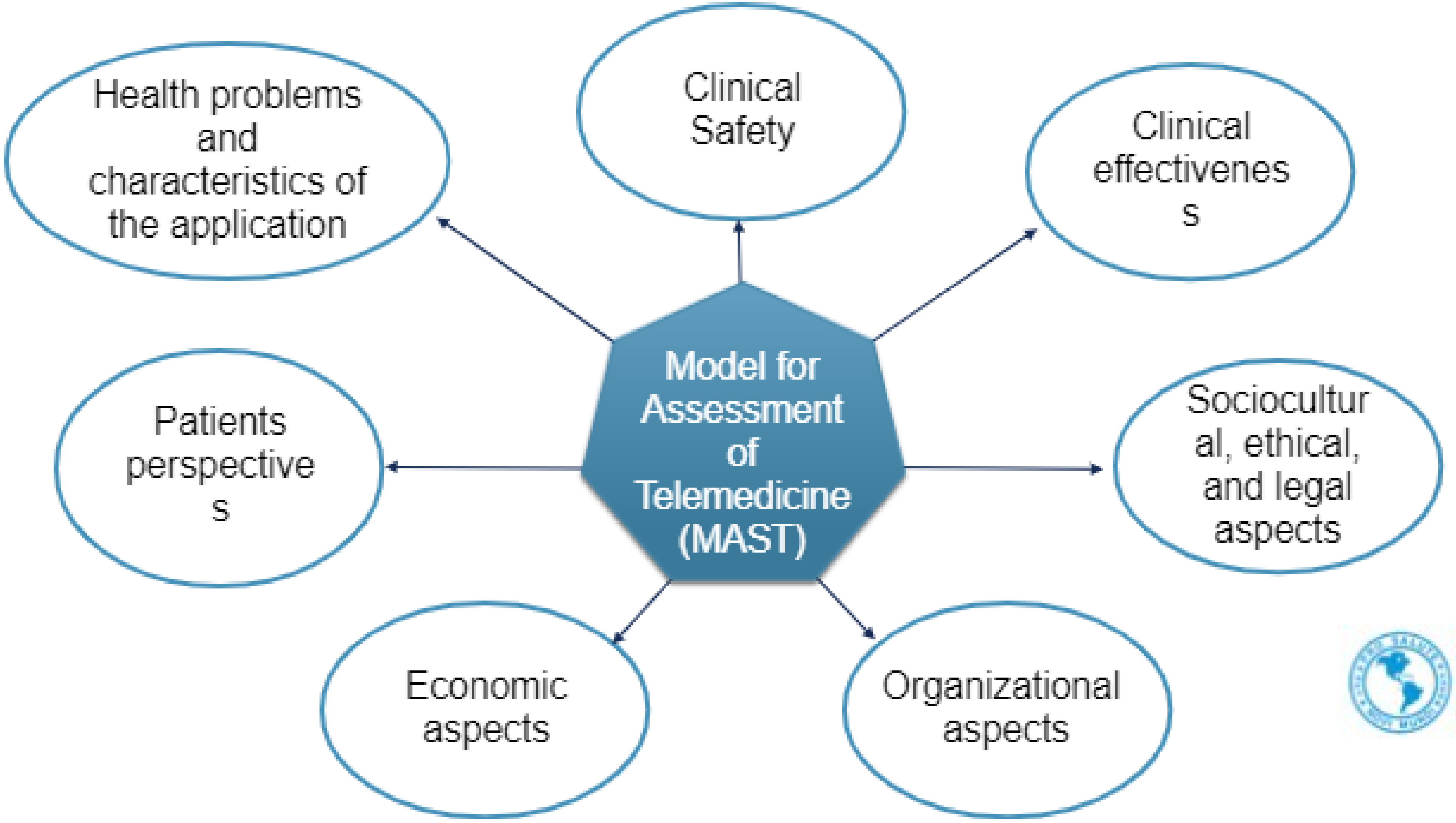


Pan American Health Organization

1. Are there any physical, mental health, or social health **barriers** that might affect participation or engagement with online CBE?

2. What about **facilitators** that might help engagement with online CBE?

3. What are your thoughts on online CBE and its ability to impact disability (I.e. symptom management, wellbeing)? What about healthy aging?

4. What are some technical factors that may affect the viability of an online CBE program? (reliability of internet, digital literacy, access to technology)

5. What content should an online CBE program include, in order to address the needs of clients/patients?

6. What platforms do you use for tele-rehabilitation/tele-coaching/online exercise/online service delivery?  (Web-based platforms, teleconferences, smart-phone apps, etc)

7. What are your thoughts regarding data-tracking for motivation and monitoring progress (e.g. using wireless activity monitors like apps or a Fitbit to motivate and measure physical activity)?

8. Can you speak about methods used by organizations providing the online programs, as well as the users, to ensure privacy and confidentiality?

9. Do users generally feel *physically* safe while using online exercise programs?

10. How do you feel end-users **trust** using online platforms for exercise programs?

11. Are there any personal or intrinsic factors that might affect an end-users **access** to a tele-coaching based program?

12. We know that the social and physical environment are important to accessing exercise programs, especially considering the recent COVID-19 pandemic. Are there any environmental factors that may affect **access** to online CBE?

13. Can you speak to any social factors that might affect **access** to online CBE?

### Break?

14. Are end-users able to advocate for themselves if they feel their needs are not being met through an online platform?

15. Can you think of any **ethical** consequences of widespread implementation of online CBE programs?

16. **Persons Living with HIV**: Can you speak more about the stigma surrounding HIV and how that might affect your willingness to participate in or engage with an online CBE intervention?

17. How might the cost of participating in/delivering an online CBE program differ compared to in-person programs?

18. What is the feasibility of providing CBE online compared to in-person?

19. In your experience, what is the best way to monitor the success of an online CBE program (or a similar online exercise program)?

20. Can you speak about the culture surrounding the use of online platforms for health services and exercise delivery?

21. Can you speak about your experience (good and bad) during the process of switching from in-person to online services?

22. What resources do you think would be needed for implementation of an online CBE?

### Section 3

RECOMMENDATIONS

23. What do you think are some overall recommendations that will be important for developing and implementing a future online CBE program for PLWH?

### Thank-you!!
