## Supplemental File 3 for "Considerations for Developing and Implementing an Online Community-Based Exercise Intervention for Adults Living with HIV: a qualitative study"

***Human Participant Ethics Protocol Submission***
***University of Toronto***

*CONFIDENTIAL*

### 0 - Identification

**RIS Human Protocol Number**
**39603**

**Protocol Title**
Tele-coaching in HIV and Exercise: Considerations for Developing and Implementing an Online Community-Based Exercise (CBE) Intervention for Adults Living with HIV

**Protocol Type**
**Investigator Submission**

#### Applicant Information

**Applicant Name**
**Kelly O'Brien**

**Rank / Position**
Assoc Professor

**Department / Faculty**
Dept of Physical Therapy - Faculty of Medicine

**Business Telephone**
416-978-0565

**Extension**

**Email Address**


#### Collaborators/Co-Investigators

| Name | Department | Email | Designation | Alt Contact |
|---|---|---|---|---|
| Julia Kobylianski | Department of Physical Therapy | | Co-Investigator & Alt | X |
| Bernice Lau | Department of Physical Therapy | | Co-Investigator & Alt | X |
| Sukhbir Manku | Department of Physical Therapy | | Co-Investigator & Alt | X |
| Isha Sharma | Department of Physical Therapy | | Co-Investigator & Alt | X |
| Anna Wong | Department of Physical Therapy | | Co-Investigator & Alt | X |
| Soo Chan Carusone | Casey House | | Co-Advisor | |
| Francisco Ibanez-Carrasco | Dalla Lana School of Public Health | | Co-Advisor | |

#### Projected Project Dates

Estimated Start Date
1-Sep-20

Estimated End Date
31-Aug-21

### 2 - Location

**Location of the Research:** ☒ University of Toronto ☐ Other Locations

#### Administrative Approval/Consent



**OFFICE OF RESEARCH ETHICS**
McMurrich Building, 12 Queen’s Park Crescent West, 2nd Floor, Toronto, ON M5S 1S8 Canada
Tel: +1 416 946-3273 ● Fax: +1 416 946-5763 ● ● http://www.research.utoronto.ca/for-researchers-administrators/ethics

Administrative Approval/Consent Needed: ( ) Yes (•) No

Community Based Particatory Research Project? ( ) Yes (•) No

**Other Ethic Boards Approval(s)**

Another Institution or Site involved? ( ) Yes (•) No

### 3 - Agreements and Reviews

#### Funding

Project Funded? (•) Yes ( ) No

**Internal U of T Funding**

| Source | Status | Peer Reviewed |
|---|---|---|
| Connaught Fund and Ontario HIV Treatment Network | Awarded | X |

#### Agreements

Funding/non-funding Agreement in Place? ( ) Yes (•) No

Any Team Member Declared Conflict of Interest? ( ) Yes (•) No

#### Reviews

[X] This research has gone under scholarly review by thesis committee, departmental review committee, peer review committee, or some other equivalent

Type of Review : -e.g.: departmental research committee, supervisor, CIHR, SSHRC, OHTN, etc.

This research protocol has undergone three reviews / revisions by the advisory committee and the protocol was presented to faculty evaluators external to t

[X] This review was specific to this protocol

[ ] The review was part of a larger grant

[ ] This research will go under scholarly review prior to funding

[ ] This review will not go under a scholarly review

### 4 - Potential Conflicts

**Conflict of Interest**

Will researchers, research team members, or immediate family members receive any personal benefit? ( ) Yes (•) No

**Restrictions on Information**

Are there any restrictions regarding access to, or disclosure of information (during or after closure)? ( ) Yes (•) No

**Researcher Relationships**

Are there any pre-existing relationships between the researchers and the researched? (•) Yes ( ) No

Relationship Description

Participants may include persons whom Kelly O’Brien, Soo Chan Carusone, or Francisco Ibanez-Carrasco (supervisors) have worked with as a colleague in a community-based research or educational capacity. Student investigators, who have no relationship with the potential participant, will recruit and obtain consent. Interested participants will be invited to contact the study student co-investigators, who have no relationship with the potential participants, who will email information and discuss the study in detail and if applicable, will obtain consent.



**OFFICE OF RESEARCH ETHICS**
McMurrich Building, 12 Queen’s Park Crescent West, 2nd Floor, Toronto, ON M5S 1S8 Canada
Tel: +1 416 946-3273 ● Fax: +1 416 946-5763 ● ● http://www.research.utoronto.ca/for-researchers-administrators/ethics

**Collaborative Decision Making**

Is this a community based project - i.e.: a collaboration between the university and a community group? ○ Yes ◉ No

### 5 - Project Details

**Summary**

**Rationale**

Describe the purpose and scholarly rationale for the project

RATIONALE

The advent of antiretroviral therapy (ART) has dramatically decreased the mortality and morbidity of people living with HIV (PLWH) (1). However, due to a combination of HIV, side-effects of ART, aging, and multimorbidity, adults living with HIV can face multiple complex and potentially episodic physical, cognitive, mental, and social health-related difficulties, which can be conceptualized as disability(2–5).

Exercise is an effective strategy to improve disability and overall well-being of adults living with HIV, however, a majority of adults living with HIV do not engage in regular exercise due to barriers such as stigma, lower socioeconomic status (SES), physical symptoms, and transportation accessibility.(6–10). Community-based exercise (CBE) can promote social support, encourage regular exercise, improve physical function, and facilitate self-management strategies in adults living with HIV (11,12 ). Despite the benefits of CBE, adults living with HIV may experience barriers which limit their participation, including discomfort with conventional gym environments, social, financial, and geographical barriers, self-image issues, stigma, and difficulty initiating exercise following periods of inactivity (9).

Tele-rehabilitation is the delivery of rehabilitation programs or services via technology, and is a potential strategy to increase accessibility to, and adherence with rehabilitation programs (13–15 ). Tele-rehabilitation can mitigate barriers, such as transportation, financial, and time commitment, and has potential to improve access to exercise programs for adults living with HIV (16).

Tele-coaching, a component of tele-rehabilitation that involves the use of technology for remote supervision, guidance, and communication of an exercise program, can help improve physical activity levels and manage symptoms for people living with chronic diseases (17–20). This approach may be particularly relevant to address additional disability experienced by people living with HIV, as well as the broader population, as a result of physical distancing measures and closures of fitness facilities amid the current COVID-19 pandemic (21,22). Nevertheless, a paucity of evidence exists on the implementation of remote CBE programming for adults living with HIV (9,11–15).

PURPOSE and OBJECTIVES:

The purpose of our study is to describe considerations for developing and implementing an online CBE intervention for adults living with HIV, from the perspectives of key stakeholders with a role in online CBE implementation for adults living with HIV including: persons living with HIV, rehabilitation or other healthcare professionals, fitness professionals, eLearning educators, and representatives from AIDS Service Organizations (ASOs).

Specific objectives are:

[1] To describe the need for, and utility of, online CBE interventions with adults living with HIV and

[2] To identify the key factors to consider in developing and implementing an online CBE intervention for adults living with HIV.

Results will yield recommendations to inform the future development and implementation of online CBE programming tailored to increase accessibility to engaging in physical activity for adults living with HIV.





**OFFICE OF RESEARCH ETHICS**
McMurrich Building, 12 Queen’s Park Crescent West, 2nd Floor, Toronto, ON M5S 1S8 Canada
Tel: +1 416 946-3273 ● Fax: +1 416 946-5763 ● ● http://www.research.utoronto.ca/for-researchers-administrators/ethics

feasibility study. Mult Scler Relat Disord. 2018;22:90-96. doi:10.1016/j.msard.2018.03.020
20. Lai B, Bond K, Kim Y, Barstow B, Jovanov E, Bickel CS. Exploring the uptake and implementation of tele-monitored home-exercise programmes in adults with Parkinson's disease: A mixed-methods pilot study. J Telemed Telecare. 2020;26(1-2):53-63. doi:10.1177/1357633X18794315
21. Shiau S, Krause KD, Valera P, Swaminathan S, Halkitis PN. The Burden of COVID-19 in People Living with HIV: A Syndemic Perspective. AIDS Behav. April 2020. doi:10.1007/s10461-020-02871-9
22. Marziali ME, Card KG, McLinden T, Wang L, Trigg J, Hogg RS. Physical Distancing in COVID-19 May Exacerbate Experiences of Social Isolation among People Living with HIV. AIDS Behav. April 2020. doi:10.1007/s10461-020-02872-8

### Methods

Describe formal/informal procedures to be used

STUDY DESIGN:
We will conduct a cross-sectional qualitative descriptive study using semi-structured one-on-one interviews. A qualitative study will enable us to use an exploratory approach, therefore capturing a breadth of perspectives as well as provide a foundational approach to addressing the limited knowledge surrounding the intersection of online CBE and the HIV community.

DATA COLLECTION:
We will conduct semi-structured interviews, using tailored interview guides with open-ended questions specific to each stakeholder group. Interviews will take place through online teleconferencing software, Zoom, and will be conducted by one interviewer and one note taker. We estimate each interview to be 30-60 minutes in duration. Interviews will be audio recorded for later verbatim transcription. We believe one-one interviews are the best method to achieve our study objectives as this method allows for participants to share personal experiences, opinions, and stories in a conversational and comfortable environment.

Interview Guide: We will use a semi-structured interview guide tailored for each stakeholder group developed based on our literature review and interview guides used in similar studies (Appendix D). We will begin with a preamble, introducing the interviewer and note taker to the participant, and providing an overview of the interview process.

Demographic Data: The interviewer will ask demographic questions to gather information about 1) age (in years), 2) gender, 3) stakeholder group(s) the participant identifies with, 4) experiences working with people living with HIV (PLWH) (yes/no), and 5) experiences with tele-health/tele-rehabilitation/tele-coaching/exercise online for PLWH (yes/no). We will use responses to (3) to select the most applicable probing questions for each interview.

We will start by asking all stakeholders the same question: to describe their experiences with, or interests in, online tele-health/tele-rehabilitation or tele-coaching (CBE interventions) with PLWH or other chronic conditions. We will then ask a series of open-ended questions to address our study objectives with tailoring of question wording to align with the tele-health, tele-rehabilitation, eLearning, or exercise experiences among each stakeholder group.

SECTION 1 (Objective #1): We will ask participants about the need for online CBE programs for PLWH, specifically why online CBE is an ideal option for self-management in PLWH: the barriers and gaps that need to be filled to address those to physical activity for PLWH, as well as ask about who might benefit the most, and who may not. We will then ask about the utility of online CBE interventions for each of the different stakeholder groups. Questions in this section will be the same for all stakeholder groups.

SECTION 2 (Objective #2): We will then ask participants about the factors they think are important for developing and implementing online CBE interventions for PLWH. Our probing questions are informed from the MAST framework; we will specifically ask about factors pertaining to: 1) health (individual physical, cognitive, mental-emotional, and social health factors) 2) clinical impact (impact on disability, healthy aging), 3) characteristics of the application (technical factors, types of exercise interventions, data tracking); 4) safety (safety of exercise and technical security/privacy), 5) person-specific and environmental factors (personal attribute factors, trust, capacity of ICT use, access, empowerment, and social support with online CBE), 6) sociocultural, ethical, and legal factors of online CBE (HIV stigma), 7) economic factors (cost for the PLWH, organization delivering the intervention), and 8) organizational factors (capacity for program evaluation, who will be delivering the online CBE intervention, resources needed for implementation and delivery, and culture of ICT use). Each stakeholder group may be able to speak more to specific factors more than others and we will tailor our probing questions accordingly.

SECTION 3: Finally, we will ask all stakeholders about overall recommendations they have for developing and implementing an online CBE program for PLWH.

Procedural Rigor: We will pilot the interview guides and conduct mock interviews with consenting members of the target population to improve our interview skills and refine our interview guides. Two members of the team will conduct the interviews to ensure consistency with the data collection process. One team member will conduct the interview, while the other will take field notes regarding non-verbal communication, and reflections on the interview process that will inform our refinement to the interview guide. Interviewers will discuss and record their impressions of each interview in a study log, which can then be compared to other interviews or the written transcription during analysis. We will then meet as a team throughout data collection to review the study log and refine the interview guide.

DATA ANALYSIS:
The interviews will be audio-recorded using a separate audio recording device. These audio recordings will then be transcribed verbatim by an investigator who was not present at the interview, in order to minimize potential bias. The investigators will work collaboratively to review these transcriptions and to identify a coding structure and establish themes to be used for the rest of the analysis. We will then analyze the transcribed interviews to identify codes and overarching themes, using thematic analysis.

Demographic data will be collected during the interview. We will de-identify this data, analyze it using descriptive analysis, and it will ultimately be used to better contextualize the results of the study.

To optimize anonymity, we will de-identify collected data (i.e. demographic data, transcriptions) by assigning an alpha-numeric code to each participant. Additionally, as interview transcripts can contain clues to a person's identity, we will ensure confidentiality through reporting results by referring to participants and including quotation only using their assigned alpha-numeric code, as well as avoiding using quotes that may lead to residual disclosure of individuals.

List of appendices:
1. Appendix A – Recruitment Flowchart (Date last revised: June 25, 2020)
2. Appendix B – Recruitment Email Templates (Date last revised: June 25, 2020)
3. Appendix C – Information & Consent Form (Date last revised: June 25, 2020) - REVISED AUGUST 15, 2020
4. Appendix D – Interview Guide (Date last revised: June 25, 2020)
5. Appendix E – Data Analysis Flowchart (Date last revised: June 25, 2020)
6. Appendix F – Knowledge Translation Plan (Date last revised: June 25, 2020)

Copies of questionnaires, interview guided and/or other instruments used

| Document Title | Document Date |
|---|---|
| Interview Guide | 2020-06-25 |
| Data Analysis Flow Chart | 2020-05-25 |
| Knowledge Translation Plan | 2020-06-25 |


OFFICE OF RESEARCH ETHICS
McMurrich Building, 12 Queen's Park Crescent West, 2nd Floor, Toronto, ON M5S 1S8 Canada
Tel: +1 416 946-3273 ● Fax: +1 416 946-5763 ● ● http://www.research.utoronto.ca/for-researchers-administrators/ethics

**Clinical Trials**

Is this a clinical trial? ( ) Yes (•) No

### 6 - Participants and Data

Participants and/or Data

What is the anticipated sample size of number of participants in the study? 15

Describe the participants to be recruited, or the individuals about whom personally identifiable information will be collected. List the inclusion and exclusion criteria. Where the research involves extraction or collection personally identifiable information, please describe where the information will be obtained, what it will include, and how permission to access said information is being sought.

PARTICIPANTS:
Target Population: We will target stakeholders with a role in online CBE implementation for adults living with HIV, termed “online CBE stakeholders”, with experience, expertise, or interest in areas including any combination of HIV, tele-coaching (exercise), tele-rehabilitation, or eLearning. We define ‘online CBE stakeholders’ as:

- Persons living with HIV, with experience or interest in online exercise applications or interventions;
- Rehabilitation professionals or other health care professionals with a role in the rehabilitation of people living with chronic disease with experience in tele-health or rehabilitation interventions;
- Fitness personnel or managers engaged in online exercise personal instruction or online exercise class delivery (for-profit and non-profit sectors);
- Educators with experience in eLearning in the field of rehabilitation or chronic disease management;
- Representatives from ASOs with experience delivering health or social support services remotely.

INCLUSION CRITERIA:
We will recruit English-speaking adults who self-identify as representing at least one of our target population sub-groups and able to participate in a Zoom (online) interview using their microphone and (optional) webcam.
Capturing perspectives from each of these five key stakeholder groups is essential to understand the multifactorial barriers that may affect the successful implementation of online CBE for PLWH. PLWH will provide end-user perspectives on facilitators and barriers to online CBE. Rehabilitation or other healthcare professionals should be included to offer insights about clinical impact and possible challenges with delivering rehabilitative services online; and consultation with fitness personnel or managers may identify ways to optimize exercise safety and quality. Recruiting educators with experience in eLearning will contribute insights into the logistics of online CBE implementation to best align with the end user’s learning style. Finally, inclusion of representatives from ASOs may provide key information regarding safe and secure delivery of support services with PLWH.
AUGUST 15, 2020 REVISION: There is no geographical limitation for stakeholders to participate in this study as our primary sampling method aims to identify and interview stakeholders with expertise, not necessarily stakeholders from specific locations. Stakeholders who meet our eligibility criteria may be known to advisors through their involvement in the Canada-International HIV and Rehabilitation Research Collaborative (CIHRRC). Additionally, the use of Zoom interviews facilitates stakeholder engagement and participation over this broader geographical scope.
SAMPLING:
We will use a combination of purposive and snowball sampling techniques to recruit participants representing one or more of the five key stakeholder groups. Purposive sampling allows for recruitment of individuals who are specifically positioned to share their expertise and perspectives on online CBE interventions. We will start by generating a sampling frame in consultation with advisors (O’Brien, Chan Carusone, and Ibáñez-Carrasco). We will include: individuals familiar to the research team, including research connections, colleagues and affiliations; as well as individuals who have no prior connection to the research team, but whose email contact information is accessible. At the end of each interview, we will use snowball sampling as this process of referral can broaden our sampling frame to include individuals who may be known to participants but previously not known to our advisory team, and/or those who did not have contact information.
SAMPLE SIZE ESTIMATION & JUSTIFICATION:
A sample size of 12 to 15 participants will allow our team to achieve representation, defined as $\geq$ 2 participants, from each of the five key stakeholder groups. Given the diversity of our target population, we do not expect to achieve data saturation. Nevertheless, our approach will allow us to form a foundational understanding of key considerations for online CBE necessary to achieve our study objectives from a breadth of perspectives. Past studies using qualitative descriptive methods to explore (in-person) CBE for PLWH used a similar sample size to our study, with three stakeholder groups from which they were able to recruit a minimum of two participants each. Our sampling frame developed by advisors, consisting of colleagues, educators, leaders in the fitness and HIV community, includes at least three potential participants representing each stakeholder group. Supplemented with snowball sampling, we expect to recruit 2-3 participants in each group to achieve our targeted sample of 12-15 online CBE stakeholders.

Is there any group or individual-level vulnerability related to the research that needs to be mitigated (for example, difficulty understanding consent, history of exploitation by researchers, or power differential between the researcher and the potential participant)? (•) Yes ( ) No

Participants may include persons whom members of the research team, have worked with as a colleague in a community-based research or educational capacity. Student investigators who have no relationship with the potential participant, will obtain consent. Interested participants will be invited to contact the study co-investigators or research coordinator who has no relationship with the potential participants, who will discuss the study in detail and if applicable, will obtain consent.
AUGUST 15, 2020 REVISION: While rare, HIV associated neurocognitive impairment may affect attention and information processing affecting the informed consent process. To address these potential challenges, we will assess capacity to consent prior to participation in the study. During the consent process prior to the interviews, we will ask individuals to relay their understanding of the study and what is involved in their participation. Additionally, we will encourage the participant to ask questions and discuss aspects of the study to ensure full understanding.

**Recruitment**


**OFFICE OF RESEARCH ETHICS**
McMurrich Building, 12 Queen’s Park Crescent West, 2nd Floor, Toronto, ON M5S 1S8 Canada
Tel: +1 416 946-3273 ● Fax: +1 416 946-5763 ● ● http://www.research.utoronto.ca/for-researchers-administrators/ethics

Is there recruitment of participant? (●) Yes ( ) No

Recruitment details including how, from where, and by whom

RECRUITMENT:
We will use a multi-step recruitment technique (Appendix A) that involves the following 6 steps:
Recruitment Step 1: (Initial Email #1): We will email two individuals from each of the five stakeholder groups with an initial request to participate (Appendix B.1). The email includes an overview of our study purpose, what's involved in participating (interview), and asking interested and willing individuals to respond (by email) if they wish to receive more information.
Recruitment Step 2: (Introductory Email #2): We will send an introductory email (Appendix B.2) to interested individuals reiterating the purpose of the study, eligibility criteria, overview of interview questions, and attach the information sheet and consent form (Appendix C). This email will also ask interested individuals to respond (by email) indicating their preferred interview day(s) and time(s). For individuals who do not respond within 7 days of being sent either the initial or introductory email, we will send Follow-Up Email #1 (Appendix B.4) or Follow-Up Email #2 (Appendix B.5), respectively. We will interpret a lack of response to such emails as a lack of interest and remove these individuals from the recruitment process.
Recruitment Step 3: (Interview Confirmation Email #3): We will schedule interested individuals for an interview, based on their indicated preferences of date/ time and send a confirmation email (Appendix B.3). This email consists of the interview date and time, instructions for connecting to the Zoom interview (password, link, etc), and we will re-attach the information and consent form indicating that we will ask for / confirm verbal consent at the start of the interview.
Recruitment Step 4: (Interview Reminder Email): We will email individuals a reminder (Appendix B.6) one day prior to their scheduled interview with the same information as the interview confirmation email. We will also attach the interview guide (Appendix D) for review prior to the interview.
Recruitment Step 5: At the end of the interview, we will ask participants if they would be willing to share information about our study with others they know who meet the eligibility criteria of one of our targeted stakeholder groups. We will wait for such individuals to contact us and begin the recruitment process again at Recruitment Step 2. Within 7 days of interview completion, we will email participants a post-interview thank you email (Appendix B.7).
Recruitment Step 6: (Evaluating Stakeholder Representation): We will continually evaluate the sample for representation of our stakeholder groups and return to Recruitment Step 1 for a second round of purposive sampling if needed.

Is participant observation used? ( ) Yes (●) No

Will translation materials be used/required? ( ) Yes (●) No

Attach copies of all recruitment posters, flyers, letters, email text, or telephone scripts

| Document Title | Document Date |
|---|---|
| Recruitment Flow Chart | 2020-06-25 |
| Recruitment-Email-Templates | 2020-06-25 |

Compensation

Will the participants receive compensation? (●) Yes ( ) No

Type of Compensation

☐ Financial

☐ In-kind

☒ Other: Gift Card

**Compensation Justification Details**

We will provide a $30 CDN electronic gift card (E-gift card) as a token of appreciation for participation in the study. We will email the gift card to participants within 7 days post-interview.

Is there a withdrawal clause in the research procedure? (●) Yes ( ) No

**Is compensation affected when a participant withdraws?**

If the participant chooses to withdraw from the study during the interview, they will still receive the E-gift card. If the participant does not meet the eligibility criteria at the start of the interview, the interview will not continue and the participant will not receive the E-gift card.

### 7 - Investigator Experience

**Investigator Experience with this type of research**

Please provide a brief description of the previous experience for this type of research by the applicant, the research team, and any persons who will have direct contact with the applicants. If there is no previous experience, how will the applicant and research team be prepared?

This research is being done in partial fulfillment of the requirements for a MScPT degree at the University of Toronto. Members of the research team include five student researchers, one faculty advisor (O'Brien), and two co-advisors (Chan Carusone, Ibanez-Carrasco)


**OFFICE OF RESEARCH ETHICS**
McMurrich Building, 12 Queen's Park Crescent West, 2nd Floor, Toronto, ON M5S 1S8 Canada
Tel: +1 416 946-3273 ● Fax: +1 416 946-5763 ● ● http://www.research.utoronto.ca/for-researchers-administrators/ethics

i) Faculty Supervisor (Kelly O'Brien): Kelly O'Brien is a physical therapist and Associate Professor at the Department of Physical Therapy, University of Toronto. She is the lead advisor on this project. Kelly has experience conducting qualitative studies and supervising MScPT students in similar forms of research.
Co-Advisor (Soo Chan Carusone): Soo Chan Carusone is the Research Lead at Casey House and an Assistant Professor (part-time) in the Department of Clinical Epidemiology and Biostatistics, McMaster University. Soo has experience with community-based research among people living with HIV, including numerous qualitative studies. Soo and Kelly have advised 3 past MScPT projects using similar study design and data collection techniques.
Co-Advisor (Francisco Ibáñez-Carrasco): Francisco Ibáñez-Carrasco is an Assistant Professor in the Dalla Lana School of Public Health at the University of Toronto, an eLearning specialist and expert in community-based research. His research focus is on health promotion and rehabilitation for people living with HIV and other episodic and chronic conditions, online adult education, and the uses and impact of social memory. He is an AIDS activist living with HIV since 1986 and a nonfiction author specialized in autopathography. Francisco is a longstanding collaborator of Kelly and Soo and has expertise in eLearning and qualitative research.
ii) Student Researchers: Student researchers (Julia Kobylianski, Bernice Lau, Sukhbir Manku, Isha Sharma, Li Yin Wong) are new to this area of research. They will be closely advised by faculty advisors (O'Brien, Chan Carusone, Ibanez-Carrasco) throughout. To prepare to undertake this research project, students completed a literature review to become familiar with the existing research, attended lectures and completed readings on qualitative research study design, and developed the study protocol and associated Appendix documents, which was reviewed extensively and approved by the advisors. Furthermore, the students completed the TCPS2 ethics tutorial, and presented a three-minute summary of the research protocol to Daniel Gyewu, Research Ethics Officer at the University of Toronto Ethics Board in June 2020. The students developed and will continue to develop skills in qualitative research including learning about the recruitment process, developing an interview guide, understanding the steps in data collection and analysis, and considering the ethical issues associated with this research project. They will additionally engage in a data collection skills workshop; data analysis workshop; and Nvivo tutorial (September 2020 – June 2021) as part of the MScPT research curriculum. Students will also review the interview guide and practice their interview skills with a consenting member of the target population.

Are community members collecting and/or analyzing data? ○ Yes ◉ No

### 8 - Possible Risks and Benefits

#### Possible Risks

Potential Risk Details:

| | | |
|---|---|---|
| Physical Risks | ○ Yes | ◉ No |
| Psychological/emotional Risks | ◉ Yes | ○ No |
| Social Risk | ◉ Yes | ○ No |
| Legal Risk | ○ Yes | ◉ No |

Risk Description

There are no known explicit physical nor emotional risks of participating in this study, however it may be possible that certain questions may be uncomfortable for the participant to answer. There may be also be a social risk participating in this study through a Zoom interview in the home environment due to a potential lack of privacy, which may influence one's comfort level when answering interview questions, or even leading to concerns of residual HIV disclosure. The participant may choose to either skip any questions, pause the interview to take a break, or terminate the interview at any point WITH THE OPTION TO RESCHEDULE (REVISION AUGUST 15, 2020). If the participant becomes upset following the interview, they will be encouraged to follow up with their healthcare provider if needed.

#### Potential Benefits

Benefit Description

There are no direct benefits for participants in taking part in this study. However, the knowledge gained from the participant's perspectives will inform the development of a future online CBE intervention, which may in turn improve access to engaging in physical activity for adults living with HIV.

### 9 - Consent

Consent Process Details

INFORMED CONSENT
Informed Consent: Participation in the study is voluntary, and participants can refuse to answer questions or stop the interview at any time without consequence. We will inform participants of their right to withdraw consent for usage of collected data, up until 7 days after completion of the interview. Throughout the previously described recruitment process, we will provide ample information regarding the participation in this study for participants to review, and we provide a contact email and phone number should participants have questions. We will obtain and document verbal consent obtained at the start of the interview by the interviewer signing and dating the consent form, confirming that the participant has been fully informed and agrees to proceed with the study. POSSIBLE RISKS AND BENEFITS: There are no physical risks associated with participating in this study. We do not anticipate the interview questions to be sensitive in nature, nevertheless, we will direct participants to follow-up with their healthcare provider should they become upset following the interview. We will remind participants they are free to skip questions, interrupt, ask for clarification, or stop the interview at any time. As outlined in the consent form, participants will also have an option to withdraw their consent for usage of data collected up until 7 days after the completion of the interview. There may be a social risk with this study as participants engaging in a Zoom interview in the home environment with lack of privacy may potentially influence the comfort level of answering interview questions or even leading to concerns of residual HIV disclosure. We will send the interview guide in advance of the interview so participants may follow up with additional perspectives with the team via email, which we will append to the transcript as data. As such, we will recommend participants seek private spaces to conduct the interview. There are no direct benefits for participants when taking part in this study. Indirect benefits include providing information that may inform future research to improve access to engaging in physical activity for adults living with HIV.


**OFFICE OF RESEARCH ETHICS**
McMurrich Building, 12 Queen's Park Crescent West, 2nd Floor, Toronto, ON M5S 1S8 Canada
Tel: +1 416 946-3273 ● Fax: +1 416 946-5763 ● ● http://www.research.utoronto.ca/for-researchers-administrators/ethics

COMPENSATION: We will send participants a $30 electronic gift card as a token of appreciation for their involvement in the study, regardless of interview completion.
We will document verbal consent obtained at the start of the interview by the interviewer signing and dating the consent form (Appendix C, page 6), confirming that the participant...
1. Understands the information provided for the above study,
2. Has been able to consider the information, ask questions, and have had them answered thoroughly,
3. Understands that participation is voluntary and able to withdraw consent for this study up to 7 days after the completion of the interview without any penalty, Understands that the data collected during the study may be looked at by individuals from the research team, or the regulatory authorities where it is relevant, and
4. Agrees to participate in this study.

Uploaded letter/consent form(s)

| Document Title | Document Date |
|---|---|
| Information Sheet & Consent Form | 2020-06-25 |
| Appendix C - Information Sheet and Consent Form - REVISED August 15, 2020 | 2020-08-15 |

Is there additional documentation regarding consent such as screening materials, introductory letters etc.: ◯ Yes ◉ No

Uploaded letter/consent form(s)

Will any information collected in the screening process - prior to full informed consent to participate in the study - be retained for those who are later excluded or refuse to participate in the study? ◯ Yes ◉ No

Is the research taking place within a community or organization which requires formal consent be sought prior to the involvement of the individual participants ◯ Yes ◉ No

Are any participants not capable (e.g.: children) of giving competent consent? ◯ Yes ◉ No

### 10 - Debriefing and Dissemination

DeBrief

Will deception or intentional non disclosure be used? ◯ Yes ◉ No

Will a written debrief be used? ◯ Yes ◉ No

Do participants/communities have the right to withdraw their data following the debrief? ◉ Yes ◯ No

Withdrawal Process Details

As outlined in the consent form, participants will have an option to withdraw their consent for usage of data collected up until 7 days after the completion of the interview. We will remind the participant of their right to withdraw the information collected in the interview, prior to the start of the interview process. Participants may communicate their desire to withdraw their data from the study by sending an email to the research group's email address. If participants decide to withdraw from the study, their data will be removed from storage and destroyed by the research team. There will be no negative consequences for the participant in the case of withdrawal.

Information Feed Back Details following completion of a participants participation in the project

Our knowledge translation will include a process of diffusion and dissemination of information gleamed from this study (Appendix F):
Diffusion
• Students will present a poster presentation at MScPT Research Day (July/August 2021)
• Students and advisors will submit a manuscript to an open-access peer-reviewed journal (e.g. British Medical Journal Open HIV/AIDS, Disability and Rehabilitation)
• Students will present (either poster or oral presentation) at OPA and/or CPA annual congress
Dissemination
• Students will post a summary of findings on the Canada-International HIV and Rehabilitation Research Collaborative (CIHRRC) website and social media (e.g. twitter)
• Students will develop and disseminate a 1-page plain language summary, which will be provided via email to participants who selected, "I would like to receive a copy of the summary of study results by email following the completion of the study" on the consent form.
• Students will present summary briefings to stakeholders (e.g. presentations to affiliated organizations of stakeholders, such as Toronto YMCA)

Procedural details which allow participants to withdraw from the project

Participation in the study is voluntary, and participants can refuse to answer questions or stop the interview at any time without consequence. On the information sheet and consent form (Appendix C), details about participants' right to withdraw are clearly presented. If participants decide to withdraw from the study, there will be no negative consequences for the participant. We will inform participants of their right to withdraw consent for usage of collected data, up until 7 days after completion of the interview. Throughout recruitment, we will provide a contact email and phone number should participants have questions. When ensuring the capacity to consent, the participants will be asked to communicate his/her understanding of his/her right to withdraw from the study up until 7 days after the completion of the interview. The participant will also be reminded of this right if he/she appears distressed by the interview process or expresses a


**OFFICE OF RESEARCH ETHICS**
McMurrich Building, 12 Queen's Park Crescent West, 2nd Floor, Toronto, ON M5S 1S8 Canada
Tel: +1 416 946-3273 ● Fax: +1 416 946-5763 ● ● http://www.research.utoronto.ca/for-researchers-administrators/ethics

desire to stop the interview.

☐ Not Applicable

What happens to a participants data and any known consequences related to the removal of said participant

If the participant withdraws from the study, he/she may decide to remove their information from the study or allow the research team to use the information collected. The participant will have up to 7 days after the completion of the interview to request that the data collected from the interview not be used and be destroyed. If this request to revoke data is received, there will be no negative consequences for the participant.

☐ Not Applicable

List reasons why a participant can not withdraw from the project (either at all or after a certain period of time)

☒ Not Applicable

### 11 - Confidentiality and Privacy

#### Confidentiality

Is the data confidential? (●) Yes ( ) No

Will the confidentiality of the participants and/or informants be protected? (●) Yes ( ) No

List confidentiality protection procedures

We will de-identify collected data (i.e. demographic data, transcripts) by assigning an alpha-numeric code to each participant. Additionally, as interview transcripts can contain clues to a person's identity, we will ensure confidentiality through reporting results by referring to participants and including quotation only using their assigned alpha-numeric code, as well as avoiding using quotes that may lead to residual disclosure of individuals.
Demographic data will be collected during the interview. This data will be de-identified and will be used to better contextualize the characteristics of the study sample for interpretation of the results of the study.

Are there any limitations on the protection of participant confidentiality? ( ) Yes (●) No

Is participant anonymity/confidentiality not applicable to this research project? ( ) Yes (●) No

#### Data Protection

Describe how the data (including written records, video/audio recordings, artifacts and questionnaires) will be protected during the conduct of the research and subsequent dissemination of results

We will de-identify collected data (i.e. demographic data, transcripts) by assigning an alpha-numeric code to each participant. Additionally, as interview transcripts can contain clues to a person's identity, we will ensure confidentiality through reporting results by referring to participants and including quotation only using their assigned alpha-numeric code, as well as avoiding using quotes that may lead to residual disclosure of individuals.
We will ensure the use of Zoom best practices for security such as: using Zoom 5.0 (most updated version), password protected meetings, enabling the waiting room feature and locking the meeting room after the admittance of each participant. We will record the interview using a separate audio recorder.
Data Storage & Security: We will store audio recordings, and electronic files including transcripts, consent forms, contact information of participants, and study log files, in a password-protected folder on Dr. Kelly O'Brien's lab drive, at the University of Toronto. We will upload recordings to this drive and then delete the recording from the recorder immediately after each interview. To transfer files among the external advisors who do not have access to the UofT folder, we will use Sharefile; an encrypted content collaboration platform only accessible to members of the research team.

Explain for how long, where and what format (identifiable, de-identified) data will be retained. Provide details of their destruction and/or continued storage. Provide a justification if you intend to store identifiable data for an indefinite length of time. If regulatory requirements for data retention exists, please explain.

Data Storage & Security: We will store audio recordings, and electronic files including transcriptions, consent forms, contact information of participants, and study log files, in a password-protected folder on Dr. Kelly O'Brien's lab drive, at the University of Toronto. We will upload recordings to this drive and then delete the recording from the recorder immediately after each interview. To transfer files among the external advisors who do not have access to the UofT folder, we will use Sharefile; an encrypted content collaboration platform only accessible to members of the research team. Audio recordings will be deleted after manuscript publication and interview transcripts will be destroyed 5 years after study publication by Dr. Kelly O'Brien.

Will the data be shared with other researchers or users? ( ) Yes (●) No

### 12 - Level of Risk and Research Ethics Board

#### Level of Risk for the Project

Group Vulnerability: Medium


**OFFICE OF RESEARCH ETHICS**
McMurrich Building, 12 Queen's Park Crescent West, 2nd Floor, Toronto, ON M5S 1S8 Canada
Tel: +1 416 946-3273 ● Fax: +1 416 946-5763 ● ● http://www.research.utoronto.ca/for-researchers-administrators/ethics

Research Risk: Low

Risk Level: 1

Explanation/Justification

Explanation/Justification detail for the group vulnerabilty and research risk listed above

Medium group vulnerability potentially for some participants based on past experiences and backgrounds of target population, including people living with HIV, who may have experienced increased stigma and increased social risk due to potential disclosure of sensitive personal information.
We anticipate low research risk as, due to the nature of this study, participants will not be subject to any physical, emotional, cognitive risk throughout the research process. For instance, the interview process will comprise of questions regarding the participant's experience with, or knowledge of community-based exercise programs, online exercise programs, potential needs for, and utility of community-based exercise programs, and any key factors (such as barriers and facilitators) to consider when developing or implementing an online community-based exercise program. These questions are not sensitive in nature, but rather aim to understand the knowledge and experiences of the participants.

Research Ethics Board

REB Associated with this project: HIV

### 13 - Application Documents Summary

Uploaded Documents

| Document Title | Document Date |
|---|---|
| Cover Letter - Response to REB Reviews Summary | 2020-08-16 |
| Interview Guide | 2020-06-25 |
| Data Analysis Flow Chart | 2020-05-25 |
| Knowledge Translation Plan | 2020-06-25 |
| Recruitment Flow Chart | 2020-06-25 |
| Recruitment-Email-Templates | 2020-06-25 |
| Information Sheet & Consent Form | 2020-06-25 |
| Appendix C - Information Sheet and Consent Form - REVISED August 15, 2020 | 2020-08-15 |

### 14 - Applicant Undertaking

I confirm that I am aware of, understand, and will comply with all relevant laws governing the collection and use of personal identifiable information is research. I understand that for research involving extraction or collection of personally identifiable information, provincial, federal, and/or international laws may apply and that any apparent mishandling of said personally identifiable information, must be reported to the office of research ethics.

As the Principal Investigator of the project, I confirm that I will ensure that all procedures performed in accordance with all relevant university, provincial, national, and/or international policies and regulations that govern research with human participants. I understand that if there is any significant deviation in the project as originally approved, I must submit an amendment to the Research Ethics Board for approval prior to implementing any change.

☒ I have read and agree to the above conditions


**OFFICE OF RESEARCH ETHICS**
McMurrich Building, 12 Queen's Park Crescent West, 2nd Floor, Toronto, ON M5S 1S8 Canada
Tel: +1 416 946-3273 ● Fax: +1 416 946-5763 ● ● http://www.research.utoronto.ca/for-researchers-administrators/ethics

RIS Protocol Number: 39603

Approval Date: 17-Aug-20

PI Name: Kelly O'Brien

Division Name:

Dear Kelly O'Brien:

Re: Your research protocol application entitled, “Tele-coaching in HIV and Exercise: Considerations for Developing and Implementing an Online Community-Based Exercise (CBE) Intervention for Adults Living with HIV”

The HIV REB has conducted a Delegated review of your application and has granted approval to the attached protocol for the period 2020-08-17 to 2021-08-09.

If this research involves face-to-face (F2F) in person research, please note that REB approval alone is not sufficient to commence research. You must wait for an approval letter from the F2F COVID-19 Review Committee. The approval letter will be sent to the Principal Investigator's email address once the Committee has deemed the F2F in-person research ready to start.

Please be reminded of the following points:

An **Amendment** must be submitted to the REB for any proposed changes to the approved protocol. The amended protocol must be reviewed and approved by the REB prior to implementation of the changes.

An annual **Renewal** must be submitted for ongoing research. Renewals should be submitted between 15 and 30 days prior to the current expiry date.

A **Protocol Deviation Report** (PDR) should be submitted when there is any departure from the REB-approved ethics review application form that has occurred without prior approval from the REB (e.g., changes to the study procedures, consent process, data protection measures). The submission of this form does not necessarily indicate wrong-doing; however follow-up procedures may be required.

An **Adverse Events Report (AER)** must be submitted when adverse or unanticipated events occur to participants in the course of the research process.

A **Protocol Completion Report** (PCR) is required when research using the protocol has been completed. For ongoing research, a PCR on the protocol will be required after 7 years, (Original and 6 Renewals). A continuation of work beyond 7 years will require the creation of a new protocol.

If your research is funded by a third party, please contact the assigned Research Funding Officer in Research Services to ensure that your funds are released.

Best wishes for the successful completion of your research.



OFFICE OF RESEARCH ETHICS
McMurrich Building, 12 Queen’s Park Crescent West, 2nd Floor, Toronto, ON M5S 1S8 Canada
Tel: +1 416 946-3273 ● Fax: +1 416 946-5763 ● ● http://www.research.utoronto.ca/for-researchers-administrators/ethics
